# Intellectual disability and cerebellar hypoplasia in autism reported by associative learning

**DOI:** 10.1101/2021.03.26.21254368

**Authors:** John P. Welsh, Jeffrey Munson, Tanya St. John, Christina N. Meehan, Elise N. Tran, K. Kawena Begay, Stephen R. Dager, Annette M. Estes

## Abstract

**Objective:** To determine how impairments in associative learning in autism spectrum disorder (ASD) relate to intellectual disability (ID) and early-childhood cerebellar hypoplasia.

**Methods:** Trace and long-delay eye blink conditioning (EBC) were performed in 62 children age 11.2 years having: 1) ASD with ID (ASD+ID); 2) ASD without ID (ASD-noID); or 3) typical development (TD). The sub-second timing of conditioned eye-blink responses (CRs) acquired to a tone paired with a corneal air puff was related to brain structure at age 2 years and clinical measures across ages 2-12 years. Because CR timing is influenced strongly by cerebellar function, EBC was used to test hypotheses relating cerebellar hypoplasia to ASD.

**Results:** Children with ASD+ID showed early-onset CRs during trace EBC that were related to early-childhood hypoplasia of the cerebellum but not of the cerebral cortex, hippocampus, or amygdala. Children with ASD-noID showed early-onset CRs only during long-delay EBC without cerebellar hypoplasia. Using EBC measures, logistic regression detected ASD with 81% sensitivity and 79% specificity while linear discriminant analysis separated ASD subgroups based on ID but not ASD severity. MRI of additional 2-year-olds with ASD indicated that early-onset CRs during trace EBC revealed ASD+ID more readily than cerebellar hypoplasia, *per se*.

**Conclusions:** Early-childhood cerebellar hypoplasia occurs in children with ASD+ID that demonstrate early-onset CRs during trace EBC. Trace EBC reveals the relationship between cerebellar hypoplasia and ASD+ID likely by engaging cerebro-cerebellar circuits involved in intellect. We emphasize that the cerebellum optimizes sensory-motor processing at sub-second intervals, impairments of which may contribute to ID.

## INTRODUCTION

Autism spectrum disorder (ASD) is a neurodevelopmental disorder with core symptoms of restricted interests, repetitive behaviors, and impairments in social communication.^1^ The severity of ASD core symptoms is highly heterogeneous and can co-occur with intellectual disability (ID), in part due to heterogeneity in the genetic and environmental influences upon the disorder. Quantifiable changes in brain morphology are promising markers of the heterogeneity in symptom severity in ASD.^2^ Early identification of individualized changes in brain development may be useful for individualizing interventions to modify the progression of ASD-related disability.

Cerebellar hypoplasia was an early documented change in brain morphology in ASD,^3,4^ although its meaning remains elusive. Alterations in cerebellar development may contribute to ASD heterogeneity, extrapolating from contemporary human studies demonstrating the involvement of certain cerebellar lobules in cognition.^5-8^ Yet, cerebellar hypoplasia with ASD has not been uniformly replicated,^9-12^ perhaps reflecting participant heterogeneity across studies.^13^ Classical eye blink conditioning (EBC) is a family of associative learning paradigms in which the sub-second timing of conditioned responses (CRs) is impaired by cerebellar dysfunction.^14-17^ Three behavioral studies have indicated that EBC may help resolve the heterogeneity of cerebellar involvement in ASD.^18-20^

The purpose of our study was therefore to examine the relationships between EBC measures of cerebellar functioning to early-childhood cerebellar hypoplasia across the full ASD spectrum, including children with ID. We hypothesized that sub-second changes in CR timing, reporting impaired associative learning, could contribute to greater understanding of the heterogeneity in symptom severity, ID, and early-childhood cerebellar hypoplasia in ASD.

## METHODS

### Standard Protocol Approvals, Registrations, and Patient Consents

The studies were approved by the University of Washington Human Subjects Division Institutional Review Board and performed according to the Declaration of Helsinki. The guardians of the participants gave written informed consent and participants gave verbal assent when possible. All participants were free to withdraw.

### Participants, Inclusion and Exclusion Criteria

A total of 89 children with ASD or TD were studied with racial demographics reflecting the Seattle area (70% White, 11% Asian, 11% Black or Multiracial, 8% Latino). Participants were 76% male, consistent with the known ASD sex distribution. ASD diagnoses were made by licensed developmental clinical psychologists at the University of Washington Autism Center using contemporaneous diagnostic criteria (Diagnostic Statistical Manual (DSM)–IV, 2004-2010; DSM-V, 2014-2017) and were confirmed by the Autism Diagnostic Observation Schedule (ADOS). Exclusion criteria for the TD group were a previous or current ASD diagnosis, suspected ASD, ASD in a first-degree relative, or having received special education services. General exclusion criteria were diagnostic or genetic conditions associated with syndromic ASD or other ID (i.e. Fragile X, Rett, and Down syndromes) and significant motor impairment (i.e. cerebral palsy, apraxia). Inclusion required vision and hearing within the normal range. Each child participated in one or both of two studies.

### Study 1: EBC across the ASD spectrum

Study 1 involved 62 children that were assessed clinically and studied with EBC at age 11.2 ± 0.2 years (*n* = 32 ASD, *n* = 30 TD). Of that group, 31 children (*n* = 25 ASD, *n* = 6 TD) had previously participated in two published longitudinal studies of intellectual and adaptive behavior development at ages 2, 3, 4, and 6 years.^21,22^ The remaining children (*n* = 7 ASD; *n* = 24 TD) were ascertained for cross-sectional analyses and studied at age 12 years only. Intellectual ability was assessed by the Mullen Scales of Early Learning (MSEL) (ages 2-4),^23^ the Differential Abilities Scales (DAS) early years (age 6), and DAS-II (age 12).^24^ ASD severity was assessed by the ADOS-Western Psychological Services (ADOS-WPS) (ages 2-6 years) and the ADOS-2 (age 12).^25^ Adaptive behavior was assessed using the Vineland Adaptive Behavior Scale, 2^nd^ Edition (VABS) (age 12).^26^

EBC includes paradigms in which a participant associates a tone conditioned stimulus (CS) with a unilateral corneal air-puff unconditioned stimulus (US).^27^ Before conditioning, the US elicits a reflex eye blink that is termed the unconditioned response (UR). Across a series of trials in which the CS precedes the US at a fixed CS-US interval, the participant acquires an eye blink to the CS that is termed the conditioned response (CR). CRs acquired to the CS exhibit onsets and peak amplitudes with latencies that adapt to the CS-US interval to cover the eye at the time of the US.

The EBC apparatus (SDI Inc.) recorded eye blinks *via* an infrared photo-sensor (Honeywell, HOA1405) positioned 2 cm from the right eye via a flexible rod attached to a helmet. A corneal air puff (100-ms, 34-kPa source pressure) delivered through a 3-mm inner-diameter tube beneath the photo-sensor was the unconditioned stimulus (US). A tone (1-kHz, 61-63 dB re: 20-µPa SPL) delivered through headphones (Philips SHN5500/37) was the conditioned stimulus (CS). Sensor output voltage was digitized (1-kHz), rectified, and smoothed (5-ms).

Figure 1 shows the EBC paradigms and measures. EBC occurred over 4 sessions (90 trials/session; 20 ± 5 s inter-trial interval; 14 ± 2 d between sessions) as described.^19,20^ Sessions 1 and 2 were trace EBC at a 700 ms CS-US interval (200 ms CS followed by a 500 ms silent period followed by the US). Session 3 was long-delay EBC (700 ms CS-US interval, 800 ms CS, co-terminating US). Session 4 was short-delay EBC (400 ms CS-US interval, 500 ms CS, co-terminating US). Participants sat in view of a caregiver and computer screen or tablet that displayed a silent video. We did not perform short-delay EBC for one ASD participant. Long-delay EBC data for one TD participant were not collected due to a technical issue.

**Figure 1.**
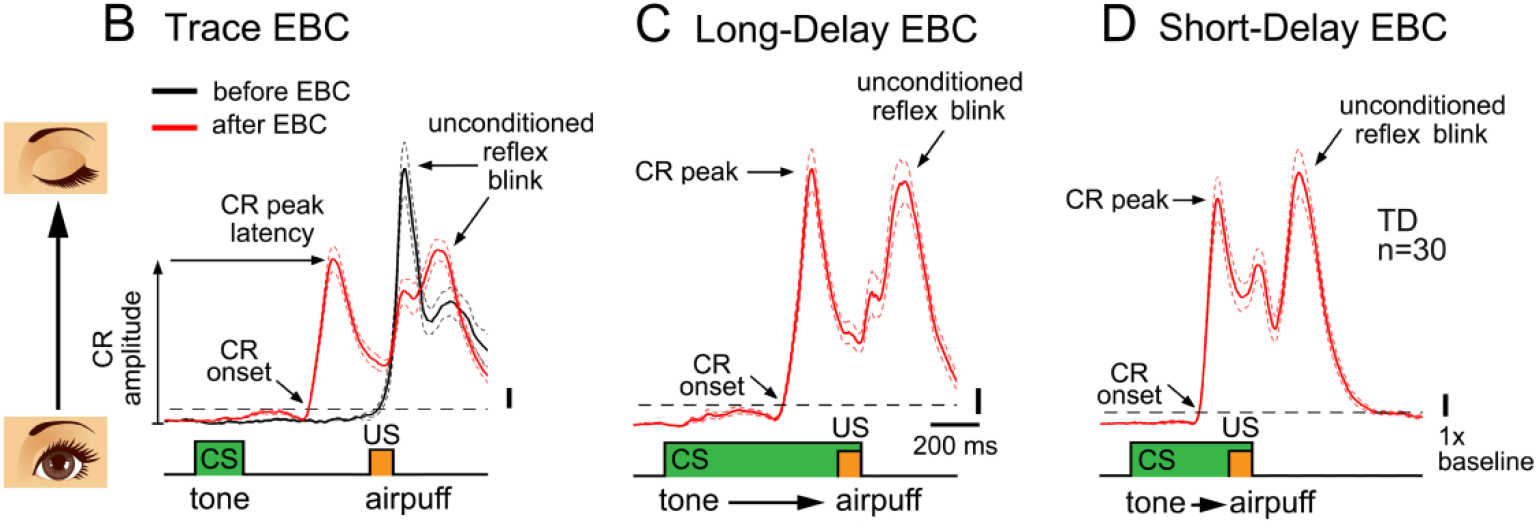
Experimental Method and EBC Paradigms. Data are mean CR topography ± SEM (dotted lines) of 30 participants with TD. (A) Image showing the setup of a school-age participant wearing headphones and a helmet that delivers the air puff nozzle and IR sensor near the right eye to elicit and measure eye blinks. The participant views a silent video during EBC. (B-C) Block diagrams (bottom) show the stimulus paradigms for trace (B), long-delay (C), and short-delay (D) EBC. Arrows indicate CR measures obtained for each participant on each session. Upward deflection in the traces represents eyelid closure.

An eye blink was defined as a deviation in photo-sensor output exceeding 10 standard deviations (SD) of a 300 ms pre-CS baseline. Eye blink onset was determined by following the photo-sensor output backward from the peak until its amplitude fell below 1 SD above the baseline. When an eye blink occurred during the CS-US interval at a latency ≥ 80 ms, it was scored as a CR. When eye blink onset was < 80 ms, it was scored as a reflex response to the tone (alpha response). Trials were excluded if peak sensor output during the baseline exceeded 5 SD of baseline, 5 SD of the mean of all baselines in the session, or if an alpha response occurred. CR amplitude was the peak photo-sensor output within the CS-US interval normalized by the pre-CS baseline.

### Study 2: Early-brain morphology with ASD+ID and early CR timing

Study 2 involved 45 children that underwent brain MRI at age 2.1 ± 0.04 years (*n* = 33 ASD, *n* = 12 TD) and were included in a published study of T2 quantitative relaxometry.^28^ Eighteen of those children participated in the 10-year longitudinal assessment and EBC experiment of Study 1 (*n* = 15 ASD, *n* = 3 TD). The remainder was a second cohort that received MRI with their age-2 diagnosis of ASD (*n* = 18) or TD (*n* = 9) contemporaneous with the Study 1 participants, but were lost to longitudinal follow-up. Of the second cohort, 12 children with ASD completed MSEL and ADOS-WPS assessments at ages 2, 3, and 4 years.

Participants with ASD were sedated using oral chloral hydrate before imaging (100 mg/kg, 2 g maximum). TD participants were scanned while asleep naturally. Axial PD and T2-weighted MRIs were acquired by a 1.5 T scanner (Signa GE, Milwaukee, WI) using a fast spin echo sequence (echo train length = 8, [effective] echo time = 13/91 ms, repetition time = 2 s, FOV = 22 cm, 256 × 160 matrix, 2.5-mm slice thickness, 0 mm gap). PD and T2 images were added to enhance gray/white contrast. T2 images were subtracted from PD images, enhancing tissue/CSF contrast. Added and subtracted images were corrected for radiofrequency inhomogeneity by homomorphic filtering. Added and subtracted images were classified using a k-means algorithm to delineate gray and white matter from the added image and brain tissue and CSF from the subtracted image.^28^ Binary images of segmented gray matter and CSF were combined using Boolean operators to exclude ventricular CSF.

Cerebral and cerebellar volumes were traced manually and volumes were calculated using a semi-automated histogram approach (MEASURE software).^28,29^ MRI tracings were performed by a trained rater blind to diagnosis and supervised by a pediatric neuroradiologist.^30,31^ Hippocampus was traced in the coronal plane (superior border: choroid fissure and inferior horn of the lateral ventricle (LV); lateral border: inferior temporal horn LV or temporal stem white matter; inferior border: parahippocampal gyrus (PHG); medial border: angle where hippocampus curved into PHG). Amygdala was traced in the axial plane (anterior border: optic nerve separation from mammillary body; lateral border: vertical line from the edge of the white matter entering amygdala; medial border: uncus; posterior border: LV temporal horn in superior slices and hippocampal head in inferior slices). Intra-rater, intra-class correlation coefficients were 0.98 and 0.96 for the right and left amygdala, respectively. Inter-rater, intra-class correlation coefficients between two raters were 0.83 for the left hippocampus and 0.92 for the left amygdala. The area of vermal lobules I–V, VI–VII, and VIII–X was measured in the midsagittal image containing the 4^th^-ventricle apex, cerebral aqueduct, and obex. Inter-rater, intra-class correlation coefficients on 10 scans for vermal lobules were: I–V, 0.97; VI–VII, 0.94; VIII–X, 0.89.

### Outcome Measures

Four CR measures were calculated for each EBC session: the percentage of trials containing a CR, mean CR onset latency, mean CR peak latency, and mean CR peak amplitude. Average CR topography was calculated for trials in which CR onset and peak occurred within 1 SD of the group mean.

Brain morphometry measures were the volumes of the cerebral cortex, cerebellum, hippocampus, and amygdala and the midsagittal area of the anterior lobe vermis, vermis lobules VI-VII, and vermis lobules VIII-X. The midsagittal areas of vermis lobules VI-X were summed to determine the area of the posterior lobe vermis.

### Statistics

Mixed-effects analysis of variance (ANOVA) was used to analyze data with repeated measures. Planned comparisons were performed with two-tailed, paired t-tests. The multivariate approaches of binary logistic regression (LR) and linear discriminant analysis (LDA) were performed using CR measures from each EBC session as independent variables. LR determined a linear equation to calculate the odds of a binary classifier variable; the natural logarithm of the calculated odds (logit) models the classifier, so that 0 logit represents equal probability of TD and ASD diagnosis given a particular set of EBC measures, while positive and negative logit represent greater probabilities of ASD diagnosis and TD, respectively. LDA derived two non-correlated sums of weighted EBC measures (canonical variables) that maximally separated 3 groups. Statistical significance of group separation was assessed by F-test. LDA classification accuracy after a significant F-test was determined using Fisher’s classification functions.^32^ Analyses were performed using SYSTAT. Statistical significance was *p* < 0.05. Data are presented as the mean ± standard error of the mean (SEM).

### Data availability

Study data are deposited at the United States National Institutes of Mental Health Data Archive. Further access will be given to investigators whose proposed use has been approved by a committee identified by the authors.

## RESULTS

### Study 1: EBC across the ASD spectrum

At age 2 years, children with ASD in the longitudinal cohort exhibited well-below-average intellectual ability (61 ± 2 standard intelligence quotient (IQ) score; 2 ± 0.2 percentile), impaired social communication, social reciprocity and social responsiveness, and a high degree of restricted interests and repetitive behaviors as compared to children in the TD group (figure 2). Over the subsequent decade, the ASD cohort separated into two groups based on their intellectual development (figure 2A,B). One group (ASD-noID; *n* = 16) exhibited gains in intellectual ability such that by age 11.9 ± 0.2 years its mean IQ (100 ± 3 standard score; 50 ± 6 percentile) approached same-age TD peers (120 ± 3 standard score; 82 ± 4 percentile). A second group (ASD+ID, *n* = 9) showed no increase in IQ (49 ± 6 standard score at 12.0 ± 0.4 years; 1 ± 0.8 percentile) and showed significant ID at the time of EBC. At age 12, the ASD-noID group demonstrated improved communication and social functioning and fewer restricted interests and repetitive behaviors (figure 2C-F).

**Figure 2.**
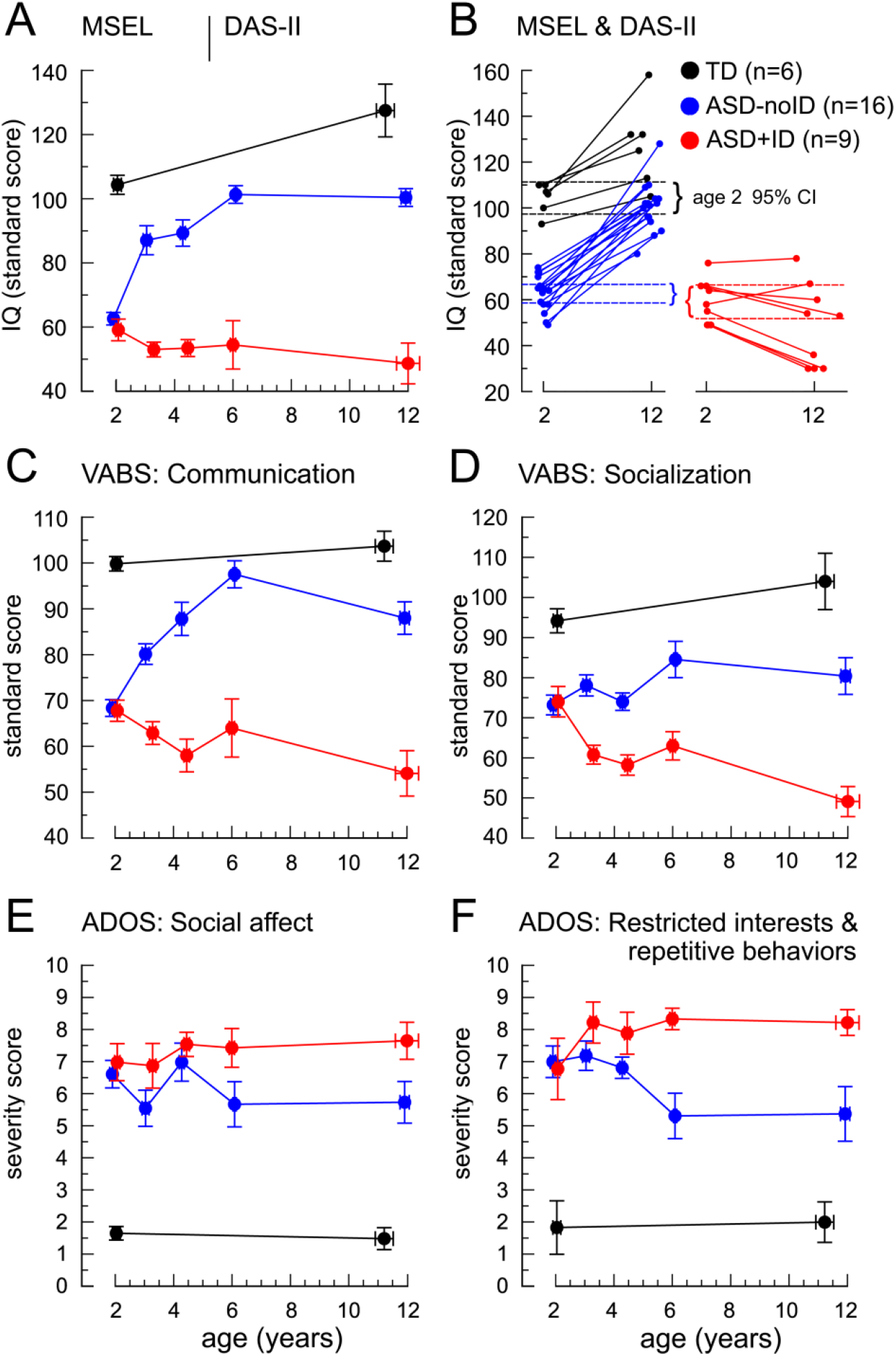
Longitudinal Characterization of Early-Childhood Intellectual and Adaptive Behavior Development for Study 1. Data in (A) and (C-F) are the mean ± SEM. Data in (B) are from individual children. (A) Standard score IQ for the longitudinal groups across ages 2-12 years. (B) Individual IQ scores at ages 2 and 12 years. (C,D) VABS standard scores for the socialization and communication domains. (E,F) ASD symptom severity scores for the social affect and restricted interest/repetitive behavior domains as measured by the ADOS.

We tested whether EBC could reveal differences in associative learning and CR timing between the ASD-noID and ASD+ID groups. At age 12 years, the longitudinal cohort was given 4 EBC sessions: 2 sessions of trace EBC, 1 session of long-delay EBC, and 1 session of short-delay EBC (figure 1). Trace EBC is defined by the presence of a stimulus-free period during the CS-US interval while delay EBC is defined by a CS that extends through the CS-US interval (figures 2B,C). There is general agreement that trace EBC elicits a more complex form of associative learning that has a greater dependence upon the telencephalon as compared to delay EBC.^33-35^ Moreover, there is agreement that the cerebellum ensures that CR timing is scaled to the CS-US interval as experimental animals and humans with cerebellar lesions cannot adapt CR timing to the CS-US interval,^14-17^ especially with lesions to the cerebellar cortex that cause early-onset CRs.^16,17^ Demonstrating the adaptation process, the onset and peak latencies of CRs acquired to a 700 ms CS-US interval were longer (figure 2C) when compared to CRs acquired to a 400 ms CS-US interval (figure 2D).

The ASD+ID group, but not the ASD-noID group, showed slower CR acquisition during trace EBC as compared to TD (*p* < 0.05; figure 3A). Additionally, the CRs of the ASD+ID group during trace EBC showed abnormally early onset (72 ± 38 ms and 1.4 ± 0.8 SD before TD) and early peak latency (73 ± 41 ms and 1.6 ± 1.0 SD before TD; both *p* < 0.01; figure 3B). Neither of those effects on CR timing was observed in the ASD-noID group.

**Figure 3.**
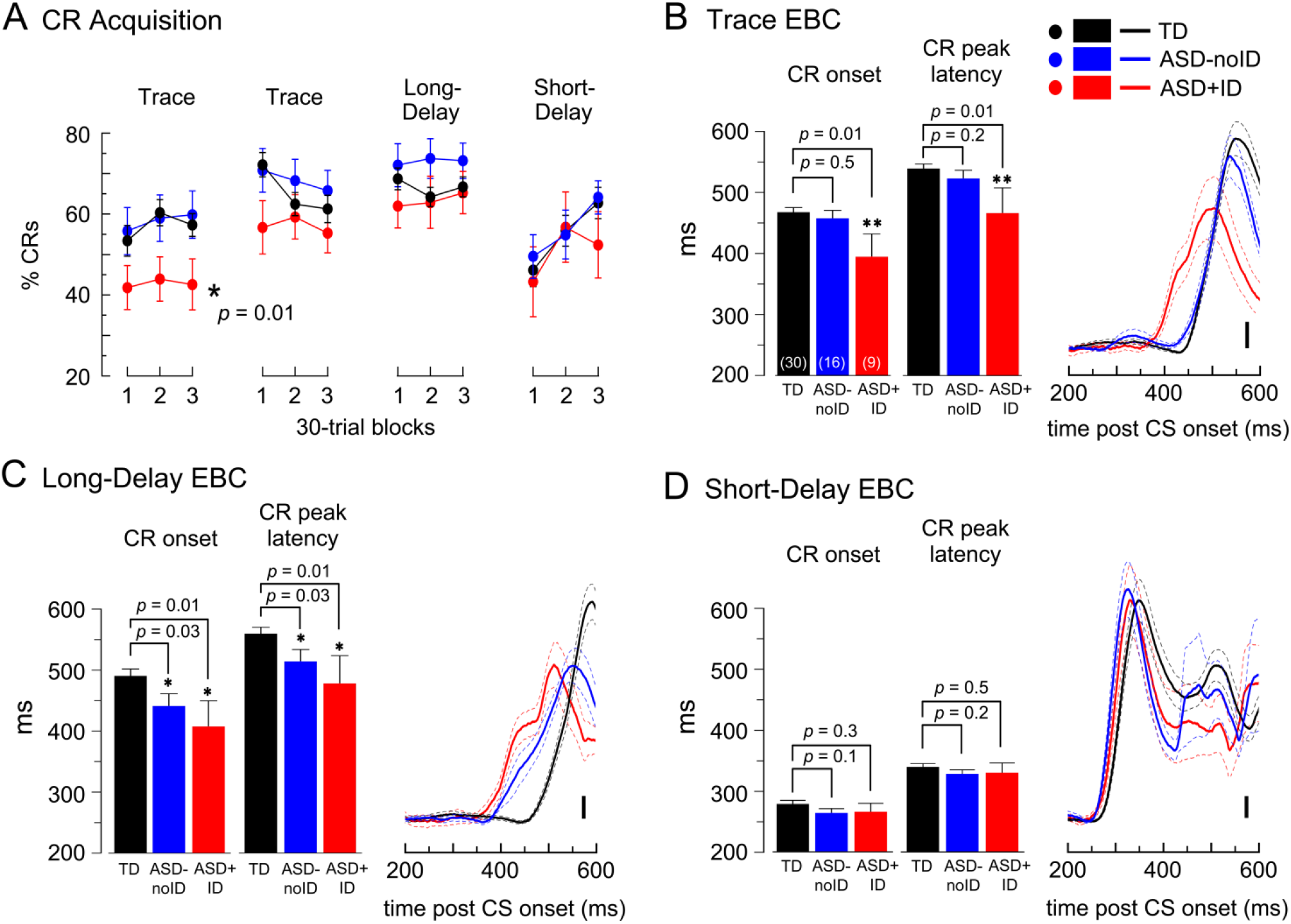
Effects of ASD-noID and ASD+ID on Trace and Delay EBC. Data are means ± SEM for the 3 groups in Study 1. (A) Percentage CRs in 30-trial blocks across 4 EBC sessions. ASD+ID differed from TD during session one of trace EBC (*F*(1,37) = 6.8, *p* = 0.013) but on no other session (all *p* > 0.1). ASD-noID did not differ from TD on any EBC session (all *p* > 0.16). (B) Mean CR onset latency, CR peak latency, and average CR topography of the 3 groups during trace EBC. (C) Mean CR onset latency, CR peak latency, and average CR topography of the 3 groups during long-delay EBC. (D) Mean CR onset latency, CR peak latency, and average CR topography of the 3 groups during short-delay EBC. * *p* < 0.05 by mixed-effects ANOVA (A) or paired t-test (B-D). Scale bar = 1x baseline (B-D).

During long-delay EBC (figure 3C), both ASD groups showed abnormally early CR onset (ASD-noID: 49 ± 20 ms, 0.8 ± 0.3 SD; ASD+ID: 82 ± 42 ms, 1.4 ± 0.7 SD before TD) and abnormally early CR peak latency (ASD-noID: 46 ± 20 ms, 0.8 ± 0.3 SD; ASD+ID: 82 ± 46 ms, 1.4 ± 0.8 SD before TD). The early-onset CRs during long-delay EBC occurred without reduction in the percentage CRs as compared to TD (figure 3A). Early-onset CRs in the ASD-noID group during long-delay EBC replicated previous reports^17-19^. During short-delay EBC, neither ASD group showed mean percentage CRs, CR onset, or CR peak latency that differed more than 0.53 ± 0.5 SD from TD (figure 3A,D).

To validate the previous analyses, we used LR to model the probability of ASD diagnosis based on EBC measures (figure 4A,B). Using 0 logit as cutoff (equal probability of ASD and TD diagnosis), EBC measures predicted ASD diagnosis correctly for 81% of cases (sensitivity) and predicted TD correctly for 79% of cases (specificity) (figure 4A). Receiver-operator characteristic (ROC) analysis produced an area under the curve of 0.87, indicating a strong positive skewing of correctly classified children when the predicted probability of ASD diagnosis based on EBC measures exceeded chance (figure 4B).

**Figure 4.**
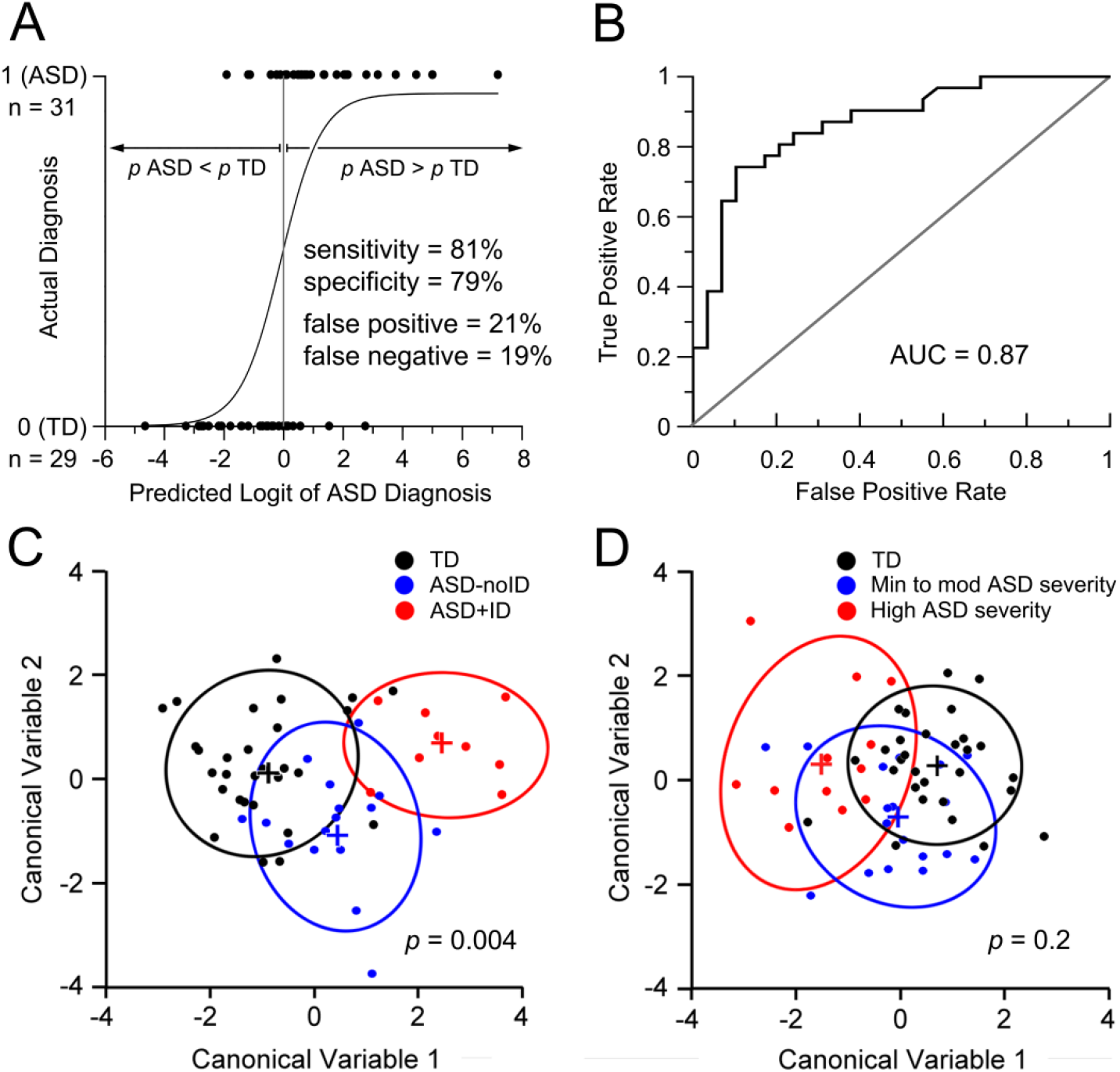
Categorization of ASD and ID by EBC measures. Data are individual cases (A,C,D), centroids (+) and 95% confidence intervals (C,D). (A) LR analysis showing individuals diagnosed with ASD or TD (dots) *vs*. the logit of the predicted probability of ASD diagnosis based on EBC measures (0 logit = chance probability of ASD or TD diagnosis). A sigmoid function is fit to the categorical data. (B) Receiver-operator curve (ROC) of the data in (A) showing 87% area under the curve (AUC), indicating robust classification of ASD and TD from EBC measures. (C) LDA of the Study 1 groups in which ASD subgroups were defined by the presence or absence of ID. LDA separated the groups with a high degree of statistical significance (*F*(2,32) = 2.2, *p* = 0.004). (D) LDA analysis of Study 1 data in which ASD subgroups were defined by ASD symptom severity (ADOS comparison scores: minimal to moderate ASD severity ≤ 6; high severity 7-10). LDA did not produce statistically-significant separation of the groups based on ASD severity (*F*(32,84) = 1,2, *p* = 0.2).

We used LDA to determine whether EBC measures can accurately classify individuals with TD, ASD-noID, and ASD+ID. LDA derived 2 canonical variables that maximally separated the 3 groups to a degree that was highly statistically significant (*p* = 0.004; figure 4C). Of note was that CR peak latencies and CR onset latencies had the four and five highest standardized weightings for canonical variables 1 and 2, respectively, indicating that differences in CR timing made the greatest contributions to group separation. Follow-up with Fisher’s classification functions categorized children as having TD, ASD-noID, or ASD+ID with 83%, 75%, and 89% accuracy, respectively.

As *post-hoc* controls, we repeated 3-group LDA after: 1) defining groups based on ASD symptom severity (minimal to moderate severity ≤ 6 *vs*. high severity, > 6 ADOS comparison score) and 2) by random assignment. LDA did not separate TD and ASD groups based on symptom severity (*p* = 0.2; figure 4D) and did not separate randomized groups (*p* = 0.96). Those analyses indicated that variation in EBC measures more closely followed ID than ASD symptom severity.

### Study 2: Early-brain morphology with ASD+ID and early CR timing

Study 2 examined the relationships among early-onset CRs, early-childhood cerebellar hypoplasia, and ID. We divided 15 children with ASD who underwent age-2 MRI and participated in Study 1 into 3 subgroups: 1) those with early CR onsets during trace EBC (ASD-early trace, *n* = 7); 2) those with early CR onsets only during long-delay EBC (ASD-early long-delay, *n* = 4); and 3) those not showing any decrease in CR onset during either paradigm (ASD-no change, *n* = 4). The mean CR onset of the ASD-early trace group was 111 ± 37 ms (2.4 ± 0.8 SD) before TD during trace EBC and 156 ± 36 ms (2.5 ± 0.6 SD) before TD during long-delay EBC. The mean CR onset of the ASD-early long-delay group was 23 ± 20 ms (0.5 ± 0.5 SD) after TD during trace EBC and 66 ± 50 ms (1.1 ± 0.8 SD) before TD during long-delay EBC. The mean CR onset of the ASD-no change group was 38 ± 12 ms (0.8 ± 0.2 SD) after TD during trace EBC and 39 ± 17 ms (0.6 ± 0.4 SD) after TD during long-delay EBC. CR topography demonstrated the differences between the ASD-early trace and ASD-early long delay groups (figure 5A,B).

**Figure 5.**
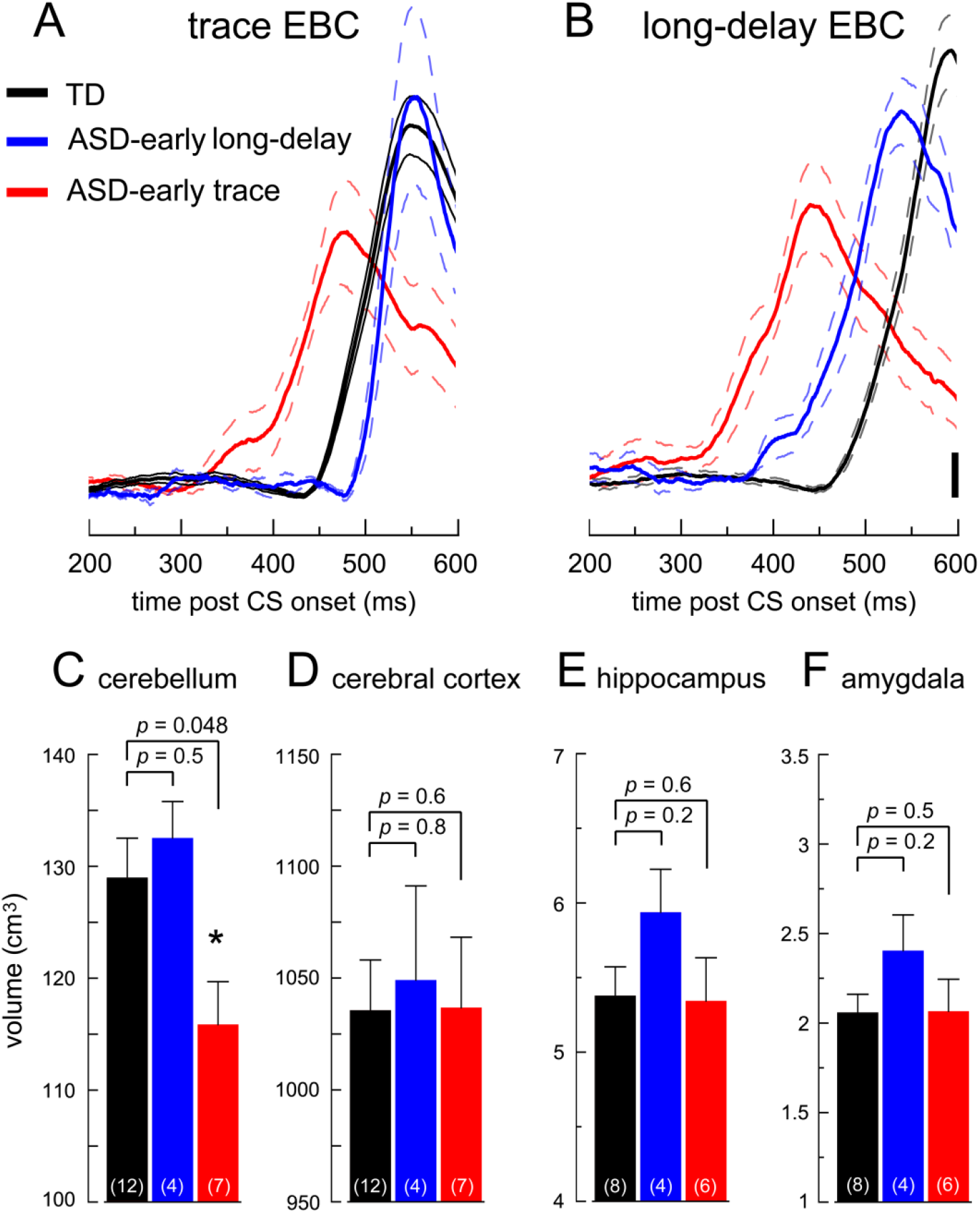
Brain Structure at Age-2 in Relation to Age-12 EBC Measures. Data are the mean ± SEM. (A,B) Mean CR topography for Study 1 participants that had brain MRI at age 2 years during trace (A) and long-delay (B) EBC. The participants are grouped by the presence of early-onset CRs during trace EBC (red, *n* = 7) or early-onset CRs only during long-delay EBC (blue, *n* = 4). (C-F) Mean volume of the cerebellum (C), cerebral cortex (D), hippocampus (E), and amygdala (F) for the groups in (A,B). **, *p* < 0.01; *, *p* < 0.05 *vs*. TD by 2-tailed, paired t-test. Scale bar = 1x baseline (A,B).

Cerebellar volume in the ASD-early trace group was significantly less than TD (118 ± 4 cm^3^ *vs*. 129 ± 4 cm^3^, 9 ± 3% decrease, *p* < 0.05). The cerebella of the ASD-early long-delay (133 ± 3 cm^3^) and ASD-no change (132 ± 3 cm^3^) groups did not differ from TD (figure 5C). Cerebellar hypoplasia in the ASD-early trace group was not accompanied by changes in the volume of the cerebral cortex, hippocampus, or amygdala (figures 5D-F).

We tested whether changes in the cerebellar anterior and posterior lobes contributed to cerebellar hypoplasia in the ASD-early trace group (figure 6A). A significant 21 ± 3% reduction in the midsagittal area of the posterior lobe vermis was found for the ASD-early trace group (*p* < 0.05; figure 6B). This change was due to a significant 21 ± 4% reduction in the midsagittal area of vermal lobules VIII-X (*p* = 0.02) and a trend-level, 20 ± 3% reduction in the midsagittal area of vermal lobules VI-VII (*p* = 0.06). The reduction in vermis area was specific to the posterior lobe, as the midsagittal area of the anterior lobe vermis in the same participants did not differ from TD (14 ± 4% reduction, *p* = 0.1). Neither the ASD-early long delay nor the ASD-no change group showed significant reductions in the area of the posterior lobe (0.6 ± 5% and 9 ± 5% reductions, respectively) or anterior lobe vermis (1 ± 4% increase, 6 ± 9% decrease, respectively) as compared to TD (all *p* > 0.05).

**Figure 6.**
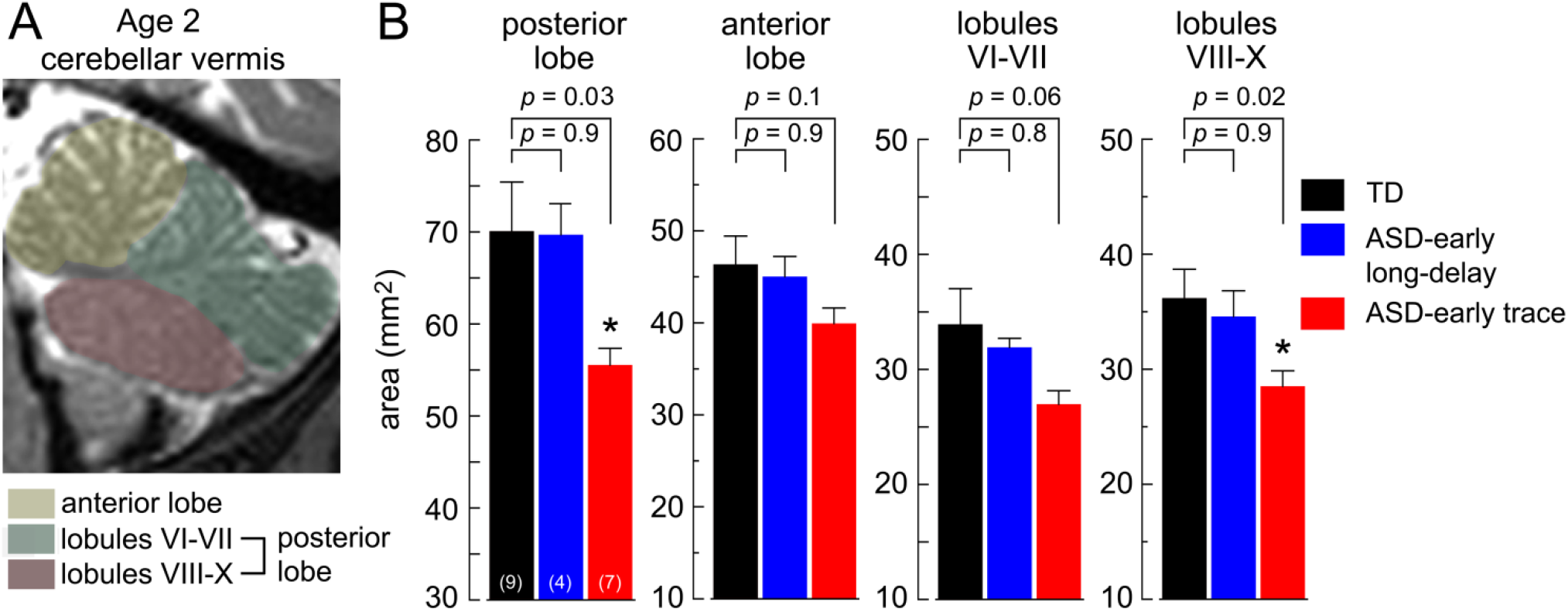
Early-Childhood Vermis Hypoplasia and CR Timing. Data are means + SEM. (A) Midsagittal image of the cerebellum at age 2 years indicating the vermal lobules studied. (B) Midline vermis area for TD (black) and ASD participants with early-onset CRs only during long-delay EBC (blue) or early-onset CRs during trace EBC (red). Participants with ASD and early-onset CRs during trace EBC showed significant reductions in lobules VIII-X and the posterior lobe vermis. Participants with ASD and early-onset CRs only during long-delay EBC did not show vermis hypoplasia. * *p* < 0.05 by 2-tailed, paired t-test.

To overcome limitations of the small sample in the previous analyses, we combined the age-2 MRI cohort from Study 1 with a second cohort of children that also had MRI at age 2 years upon diagnosis of ASD (*n* = 18) or TD (*n* = 9) but did not participate in Study 1 (combined MRI cohort *n* = 45).

With the combined MRI cohort, we tested whether posterior lobe hypoplasia was a replicable feature of reduced cerebellar volume in ASD by plotting the midsagittal area of the posterior and anterior lobe vermis *vs*. cerebellar volume (figure 7A,B). In TD, the midsagittal areas of the posterior and anterior lobe vermis scaled linearly with cerebellar volume (black, figure 7A,B). Similar slopes for the posterior and anterior lobe vermis also characterized 2-year-olds with ASD and typical-sized cerebella (blue, figure 7A,B). The scaling between vermis area and cerebellar volume was significantly altered for 2-year-olds with ASD and cerebellar hypoplasia (< 120 cm^3^; red circles, figure 7A). In those children, the posterior lobe vermis did not show a robust increase with cerebellar volume while the anterior lobe showed the scaling characteristic of TD (red, figure 7B). Quantitatively, the area of the posterior lobe vermis was nearly constant at approximately 50 mm^2^ over a 30% increase in cerebellar volume in the ASD group with cerebellar hypoplasia. The result affirmed that reduced cerebellar volume in ASD is related to hypoplasia of the posterior lobe.

**Figure 7.**
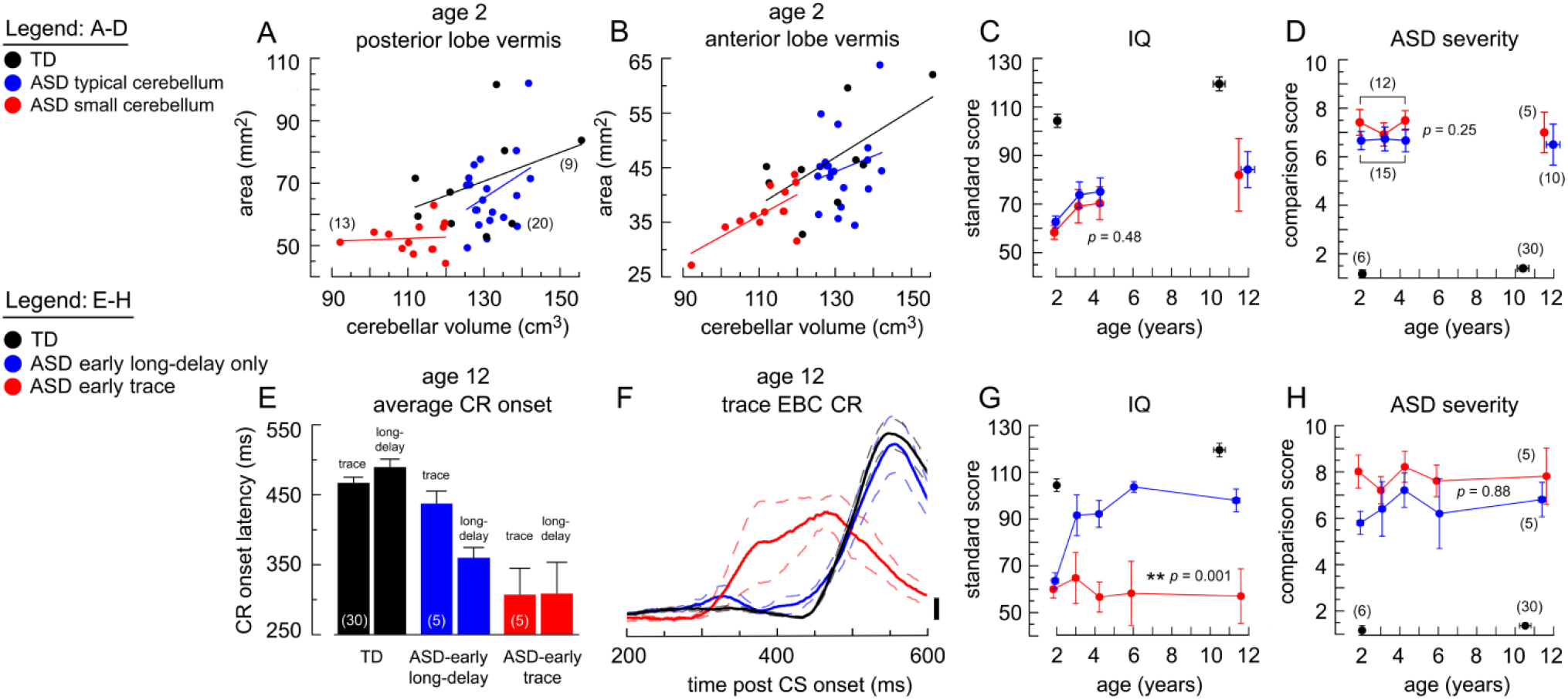
Relationships Between Age-2 Cerebellar Hypoplasia and Early-Onset CRs to ASD+ID and ASD Severity. Data are individual cases (A,B) and mean ± SEM (C-H). (A) Midsagittal area of posterior lobe vermis *vs*. total cerebellar volume for 2-year-old children (*n* = 42) in 3 groups: TD (black); ASD with cerebellar volume exceeding the ASD mean (blue); and ASD with cerebellar volume below the ASD mean (red). (B) Same individuals and groups as (A) but plotting the midsagittal area of the anterior lobe vermis *vs*. cerebellar volume. Each group is fitted with a linear regression line. (C,D) IQ (C) and ASD severity (D) for ASD groups defined by cerebellar volume normalized to the volume of the cerebral cortex above (blue) and below (red) the ASD mean as compared to TD (black). In the ASD groups, 27 children were followed longitudinally over ages 2-4. Age-12 ASD measures show the subset of children that participated at age 12. ASD groups defined by the presence or absence of cerebellar hypoplasia did not differ over ages 2-4 in IQ (effect of group: *F*(1,25) = 0.48, *p* = 0.49; group x years: *F*(2,50) = 0.004, *p* = 0.99) or ASD severity (effect of group: *F*(1,25) = 1,4, *p* = 0.25; group x years: *F*(2,50) = 0.45, *p* = 0.64). (E) CR onset latencies during trace and long-delay EBC for 12-year-old children with TD (black), ASD and early-onset CRs only during long-delay EBC (blue), and ASD and early-onset CRs during both trace and long-delay EBC (red). (F) Average CR topography during trace EBC for the groups in (E). (G,H) IQ (G) and ASD severity (H) for the groups in (E,F). Children with ASD and early-onset CRs during trace EBC showed greatly impaired intellectual development over 10 years as compared to children with ASD and early-onset CRs only during long-delay EBC (effect of group: *F*(1,8) = 10.2, *p* = 0.013; group x years: *F*(4,32) = 5.6, *p* = 0.001) but not greater ASD severity (effect of group: *F*(1,8) = 2.0, *p* = 0.19; group x years: *F*(4,32) = 0.30, *p* = 0.88). Scale bar = 1x baseline (F).

Because 27 children with ASD in the combined MRI cohort were assessed clinically over ages 2-4 years, we tested whether cerebellar hypoplasia at age 2 predisposed to less-favorable intellectual development and more severe ASD (figure 7C,D). There was no statistically significant difference in IQ (*p* > 0.4) or ASD severity (*p* > 0.2) with cerebellar hypoplasia. Of note, the ASD group with cerebellar hypoplasia showed a consistent 7% decrease in IQ over ages 2-4 compared to ASD without cerebellar hypoplasia. Because the effect size was small (Cohen’s D = 0.3), 80% power to reject the null hypothesis that cerebellar hypoplasia does not affect IQ at α = 0.05 would require groups of ∼200 participants. Thus, the significance of this potential IQ difference between the groups was minimal and supports the conclusion that cerebellar hypoplasia in ASD, *per se*, does not produce significant ID.

To better understand the relationship between ID and CR timing, we tested whether trace EBC had special relevance for ASD+ID or whether any large change in CR timing was related to ID. The former would indicate that trace EBC uniquely interrogates brain circuits impaired by ASD+ID. Thus, we compared two ASD subgroups of Study 1 participants (*n* = 5 each) that exhibited the greatest impairments in CR timing either: 1) during trace EBC (red, figure 7E-H) or 2) only during long-delay EBC (blue, figure 7E-H). For those groups, the deviation in CR onset had to precede the TD mean by more than 1.5 SD. Figures 7E,F show mean CR onsets for TD and the 2 ASD groups and mean CR topography during trace EBC. Children with ASD that had very early-onset CRs during trace EBC showed no intellectual growth over 10 years as compared to children with ASD and very early-onset CRs only during long-delay EBC who showed highly favorable intellectual growth (*p* = 0.001; figure 7G). There was no significant difference in ASD severity between the same ASD groups (*p* > 0.1, figure 7H).

## DISCUSSION

Our findings demonstrated that cerebellar hypoplasia is present in the subset of children with ASD with ID for whom early-onset CRs during trace EBC are a behavioral marker. In our prospective analysis, cerebellar hypoplasia at age 2 years did not predict ID or more severe ASD. However, in our retrospective experiment, school-aged children with ASD and early-onset CRs during trace EBC showed sustained ID throughout early childhood. Because early-onset CRs are consistent with cerebellar dysfunction, the findings are consistent with a cerebellar contribution to ASD+ID that presents in some cases as cerebellar hypoplasia at age 2 years.

The current findings help explain the meaning of cerebellar hypoplasia in ASD, an early example of altered brain morphology that helped establish ASD as a neurodevelopmental disorder.^3,4,13^ The finding of cerebellar hypoplasia in ASD has been replicated^3,4,36-39^ and has been associated with the loss of cerebellar Purkinje cells in the posterior lobe whose degenerated myelinated axons contribute to reduced cerebellar volume.^40^ Nevertheless, not all studies replicated the original observation^9-12^ and those null outcomes have been reinforced by uncertainty of how the cerebellum - traditionally viewed as a motor structure - may contribute to intellectual development or contribute to the domains of non-motor functioning impacted by ASD.

The present findings help explain cerebellar involvement in ASD by indicating that changes in implicit timing contribute to ID. Implicit timing refers to the subconscious use of time intervals that enables temporal prediction and facilitates sensory processing, cognition, prosody of speech, and movement.^41^ For instance, experience with a sub-second time interval bounded by two stimuli promotes anticipation of the second stimulus that is expressed subconsciously as an anticipatory perceptual response in the cerebral cortex and as an anticipatory motor response.^42^ The timing of CRs during EBC that adapt subconsciously to the CS-US interval with tens-of-milliseconds precision also reflects the process of implicit timing at the sub-second time-scale in which the cerebellum plays a major role.^43^ The subconscious nature of implicit timing distinguishes it from explicit timing in which conscious judgments of duration, temporal offset, and tempo can be reported verbally. As measured by human fMRI, trace EBC co-activated the cerebellum, prefrontal cortex, and hippocampus to a greater extent than delay EBC,^33^ revealing a structure of cerebro-cerebellar interaction that overlaps with the brain circuitry involved in intellect. Thus, multiple lines of evidence suggest that impairments in implicit timing contribute to ID following cerebellar pathophysiology due to a loss of sub-second precision in sensory processing and learning.

It is notable that early-onset CRs during long-delay EBC in ASD without ID have been reported by two research groups prior to our study.^18-20^ Replicating early-onset CRs during delay EBC in children with ASD builds confidence that EBC reliably distinguishes ASD from TD. However, impairments in CR timing that are restricted to delay EBC are not associated with early-childhood cerebellar hypoplasia.

By studying the full ASD spectrum, we demonstrated that only children with sustained ID showed impairments in CR timing during trace EBC. That observation indicates that cerebellar involvement in CR timing during trace and delay EBC may differ. While studies in experimental animals have implicated the posterior lobe lobule HVI as necessary for the adaptive timing of CRs during delay EBC, our study suggests that ASD with ID not only impairs the functioning of lobule HVI but may also affect postero-lateral cerebellar lobules that interact with the prefrontal cortex *via* trans-synaptic projections through the non-motor thalamus.^44^

EBC sensitively identifies children with ASD as having ID but is less sensitive for predicting ASD symptom severity. The 81% sensitivity of EBC for differentiating children with ASD from TD approximates the 88% sensitivity of a MRI approach for assessing ASD risk based on numerous gray matter measurements in neocortex.^2^ Of particular interest for our findings was that surface area expansion of the middle frontal gyrus showed the largest difference between children at low *vs*. high familial risk for ASD,^2^ within which lie Brodmann’s areas 9 and 46 that are the primary receiving zones of trans-synaptic input from cerebellar lobules^44^ that are activated during cognition.^6^ Those results indicate that EBC may be useful for detecting ID in other disorders, such as Down syndrome in which children manifest sustained ID and cerebellar hypoplasia,^45,46^ often without ASD symptoms.^47,48^

Standard clinical assessments at age 2 do not predict future intellectual development with high levels of certainty. However, combining well-understood functional tests such as EBC with brain morphology measurements in young children may enhance our ability to predict trajectories of intellectual development. Replication studies should overcome limitations of the present study by employing a prospective analysis of intellectual development and response to early behavioral intervention using coincident, early-childhood EBC and high-resolution sub-lobular measurements of cerebral and cerebellar morphology in larger cohorts than were available for the present study.

## Supporting information

STROBE

## Data Availability

https://nda.nih.gov/

## GLOSSARY

ADOS: Autism Diagnostic Observation Schedule
ASD: autism spectrum disorder
CR: conditioned response
CS: conditioned stimulus
DAS-II: Differential Ability Scales-II
EBC: eye blink conditioning
ID: intellectual disability
IQ: intelligence quotient
LDA: linear discriminant analysis
MSEL: Mullen Scales of Early Learning
SD: standard deviation
SEM: standard error of the mean
SPL: sound pressure level
TD: typical development
US: unconditioned stimulus
UR: unconditioned response
VABS: Vineland Adaptive Behavior Scales

## ACKNOWLEDGMENT

We especially thank the participating families for their generosity and unwavering commitment to brain research. We thank Dr. J. Greenson for consultation and referrals into the study, Dr. F. Reitz for technical assistance, B.F. Sparks, R. N. and Dr. D.W.W. Shaw for assistance in analyzing the brain anatomical structures, L. Tsui, C. Sinquimani for assisting with the EBC sessions, and L. Tsui, J. Raduazzo and S. Kamm for participant coordination. We gratefully acknowledge Drs. G. Dawson and S. Rogers for initiating the longitudinal study.

## STUDY FUNDING

This work was supported by a grant from the United States National Institute of Mental Health (R01 MH100887 to JPW). The diagnostic and brain imaging data of the longitudinal cohort over ages 2-6 were acquired at the University of Washington Autism and Medical Centers with the support of the United States NIMH (U54 MH066399). Many of the assessments were conducted at the University of Washington Center for Human Development and Disability, supported by the United States Eunice Kennedy Shriver National Institute of Child Health & Human Development (U54 HD083091). The funders of the study had no role in the design of the study, data collection, data analysis, data interpretation, or the writing of the report.

## DISCLOSURE

The authors report no competing interests.

## Notes

### Competing Interest Statement

The authors have declared no competing interest.

### Author Declarations

University of Washington Human Subjects IRB

